# Performance of LFSPRO *TP53* germline carrier risk predictions compared to standard genetic counseling practice on prospectively collected probands

**DOI:** 10.1101/2024.07.09.24310095

**Authors:** Jessica L. Corredor, Ruonan Li, Elissa B. Dodd-Eaton, Jacynda Casey, Ashley H. Woodson, Nam H. Nguyen, Gang Peng, Angelica M. Gutierrez, Banu K. Arun, Wenyi Wang

## Abstract

Genetic counseling and testing for germline mutations are essential for identifying individuals at increased risk for cancer. Pathogenic variants in *TP53* are diagnostic of Li-Fraumeni syndrome (LFS), a highly penetrant disorder with diverse, early-onset tumors. Current clinical guidelines, such as Chompret and Classic criteria, provide frameworks for identifying individuals at risk for likely pathogenic/pathogenic *TP53* variants; however, genetic counselors often encounter patients with features concerning for LFS that do not clearly meet established criteria, creating challenges for risk assessment and testing decisions. We evaluated whether LFSPRO, a Mendelian, family-history-based model that estimates the individual’s probability of harboring a deleterious *TP53* variant, improves carrier identification relative to guideline criteria.

In a prospectively collected cohort of 182 probands who underwent clinical genetic counseling and germline *TP53* testing, LFSPRO showed superior discrimination compared with Chompret criteria, with higher sensitivity (81% vs. 33%) and specificity (88% vs. 65%) and improved predictive values (PPV 0.53 vs. 0.14; NPV 0.96 vs. 0.85). Receiver operating characteristic analysis confirmed strong discriminatory performance (AUC=0.88). Calibration analysis using observed-to-expected ratios indicated good agreement between predicted and observed carrier frequencies (Observed/Expected=1.07).

These findings demonstrate that LFSPRO outperforms traditional guideline-based criteria for identifying *TP53* mutation carriers in real-world clinical settings. By providing quantitative, well-calibrated carrier probabilities rather than binary classifications, LFSPRO can enhance genetic counseling and support testing decisions, particularly for individuals who do not clearly meet existing criteria.

## Introduction

Li-Fraumeni syndrome (LFS [MIM: 151623]) is a rare hereditary cancer predisposition syndrome caused by germline likely pathogenic/pathogenic variants (LP/P variants) in the *TP53* (MIM: 191170), a key tumor suppressor gene that safeguards the genome from mutations^1–3^. Individuals with LFS face an increased risk of developing various cancers throughout their lifetime. The most common cancers seen in LFS families are sarcomas, breast cancer, leukemia, brain tumors, and adrenal cortical carcinoma^4,5^. The lifetime cancer risk is approximately 93% for women and 73% for men, with a 50% chance that cancers occur prior to the age of 40^6,7^. Cancer survivors in this population are at higher risk of developing additional primary cancers and treatment-related secondary cancers^7^, underscoring the importance of genetic counseling in guiding early detection, cancer risk assessment, and lifelong monitoring to better manage their health outcomes.

However, effective clinical management of LFS is challenging due to its rare nature, the overlap with other hereditary cancer syndromes, and the variable penetrance and phenotypic expression across families^8^. Although early cancer detection, as demonstrated by the 2011 comprehensive Toronto screening protocol, can dramatically improve cancer survival rates in children and adults^9^, predicting who will carry a *TP53* LP/P variant remains difficult, making it challenging to start screening at the appropriate time. Currently, risk prediction relies on clinical classification schemes such as Li-Fraumeni classic criteria^10^, Birch criteria^5^, Eeles Criteria^11^, and Chompret criteria^6,12,13^. These schemes include proband’s cancer history, age of cancer onset, and limited family cancer history in first- and second-degree relatives to guide recommendations for genetic testing for LFS. However, these criteria do not provide the detailed risk assessment often needed in genetic counseling. In cancer-focused genetic counseling sessions, genetic counselors (GCs) use risk prediction models to estimate a patient’s lifetime cancer risk, determine eligibility for surveillance and chemoprevention, and assess the likelihood of having other hereditary cancer predisposition syndromes, such as hereditary breast and ovarian cancer syndrome (MIM: 604370) and Lynch syndrome (MIM: 120435)^14^. However, such risk models did not previously exist for LFS. Given the heterogeneity in cancer presentation of LFS and the significant impact of early identification on personalized surveillance and treatment strategies, a comprehensive risk model is essential to help GCs and other healthcare providers communicate more effectively with patients.

To address this gap, Peng *et al*^15^ developed LFSPRO, a comprehensive statistical framework designed to predict both the likelihood of having a *TP53* LP/P variant and the risk of developing LFS-related cancers. Several key steps were undertaken to move LFSPRO from model development to clinical applicability (**Fig. 1a**). The development process began by identifying the clinical problem, followed by initial model development using research datasets^15^. These research datasets differ from clinically ascertained data in that they often include more detailed personal and family histories and regular follow-ups, while clinical data tends to have limited information and reflect real-world conditions. To ensure real-world applicability, the models were trained on clinical data and validated using both internal and external datasets. LFSPRO was then further validated using retrospective research-based (RB) multicenter cohorts, which provided penetrance estimates for breast cancer, sarcoma, and other cancers in LFS patients^16^. Following this, the model was further validated with a single institution’s ascertained data, which was a retrospective clinical-based (RCB) data and collected from genetic counseling sessions, reflecting real-world clinical settings with potentially less comprehensive personal and family history information compared to research datasets^17^. This clinical validation is critical for ensuring the model’s applicability in genetic counseling and clinical decision-making in real-world diverse clinical settings. Subsequently, an R Shiny app (LFSPROShiny^18^) was developed to make the model accessible to GCs and healthcare providers in a user-friendly format.

**Figure 1.**
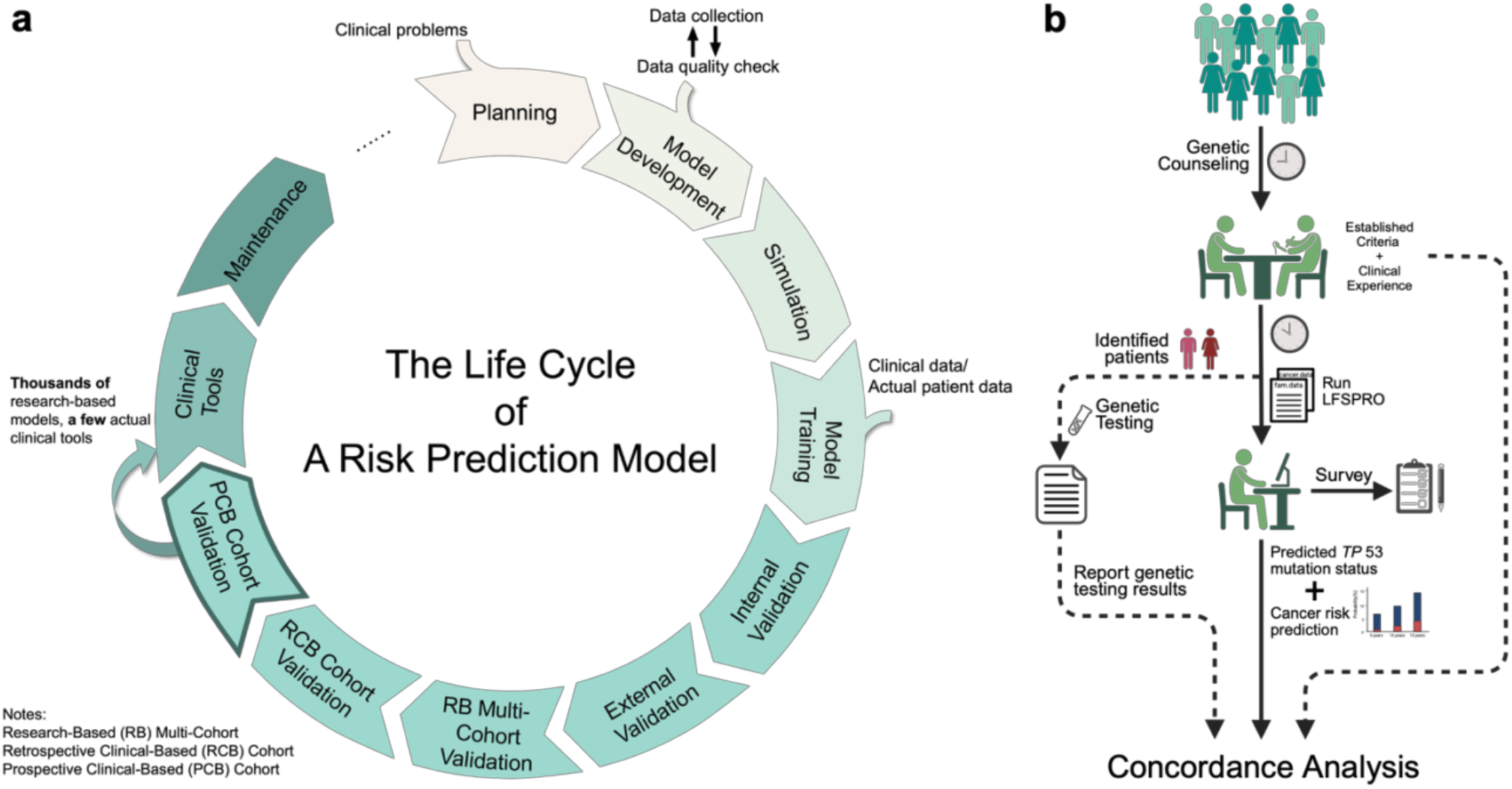
LFSPRO Development and Clinical Validation Study Design. **(a)** The systematic development and validation cycle for clinical risk prediction models, progressing from initial planning and model development through clinical tools, validation phases, and maintenance. **(b)** Patients concerning for Li-Fraumeni Syndrome (LFS) were first evaluated by genetic counselors using standard clinical criteria (e.g., Chompret or Classic) and their clinical experience. Probands identified as potentially mutation carriers underwent *TP53* genetic testing. In parallel, genetic counselors (GCs) ran the LFSPRO tool to generate *TP53* carrier probability and cancer risk predictions. GCs completed a post-session survey evaluating LFSPRO’s alignment with their clinical judgment and its potential utility. Predicted results from LFSPRO were then compared with actual genetic testing outcomes to assess concordance.

While previous studies have demonstrated LFSPRO’s strong performance in research and retrospective clinical datasets, its real-world clinical utility as a complementary tool to existing LFS risk prediction criteria remains to be fully evaluated. In this prospective study, we assessed the accuracy of LFSPRO in predicting *TP53* germline variant carrier status compared to established criteria in a real-time clinical genetic counseling setting. To further evaluate its practical value as a decision-support tool, four experienced GCs were invited to use LFSPRO following standard genetic counseling sessions with patients suspected of having LFS. Counselors reviewed the model’s output alongside their own clinical assessments and completed a structured survey providing their detailed feedback, including the model’s concordance with clinical judgment and its perceived usefulness in patient communication and decision-making.

## Methods

### Study Design and Study Population

This prospective study was designed to assess LFSPRO’s clinical utility in a real-world genetic counseling setting at the University of Texas MD Anderson Cancer Center (MDACC). Building upon previous validations that demonstrated LFSPRO’s strong performance in research-based and retrospective clinical datasets, this study aims to evaluate its performance in predicting *TP53* LP/P germline variant carrier status compared to established criteria in a prospective clinical cohort, and to assess GCs’ experience with implementing LFSPRO as part of their standard genetic counseling practice.

From December 2021 to March 2025, probands who underwent genetic counseling at MDACC and were identified as having clinical features concerning for LFS were included. Patients were eligible if they either: (1) met Classic and/or Chompret criteria for LFS testing, or (2) were deemed concerning for LFS by GCs based on their personal/family history, despite not meeting existing criteria. The latter category included individuals with early-onset LFS-spectrum tumors but small or limited family history, somatic testing suggestive of germline *TP53* variants, multiple primary cancers, and/or extensive family history of childhood and/or rare cancers.

### Data Collection

After receiving standard genetic counseling with an MDACC GC, patients identified as concerning for LFS were flagged in Progeny, which stores the internal family history database. The study team then completed a comprehensive chart review of all identified patients and assessed them using LFSPRO. Data collected included LFSPRO-predicted *TP53* LP/P variant carrier risk, whether the patient met Chompret and/or Classic criteria, decision to test, and genetic test results when available. Additional data documented for each participant included demographic information (age, sex assigned at birth), personal cancer history (type of cancer, age at diagnosis, multiple primaries), and family cancer history in first- and second-degree relatives.

Four MDACC GCs who most frequently see patients concerning for LFS were invited to use LFSPRO on their “concerning for LFS patients” after completing their standard genetic counseling session. After each instance of running LFSPRO, these counselors completed a survey regarding their experience and assessment of LFSPRO’s clinical utility for that specific patient case. A detailed illustration of the study can be seen in **Fig. 1b**. A complete survey can be seen in **Supplemental Materials.**

### Current Criteria for LFS Risk Assessment

LFS diagnosis is primarily guided by established clinical criteria designed to identify individuals with a high likelihood of carrying pathogenic *TP53* mutations. The most widely used criteria in genetic counseling include the Classic LFS criteria and the Chompret criteria (see **Table 1**). The Classic criteria, originally defined by Li and Fraumeni in 1988, have high specificity (91.0-98.1%) but low sensitivity (25.0-40.0%), making them the most stringent approach^19^. Chompret criteria were developed by Chompret et al and revisited in 2009 and 2015. The revised Chompret criteria offer improved sensitivity (83%) while maintaining reasonable specificity (47%)^13^. These criteria guide GCs in evaluating a proband’s personal cancer history, age of cancer onset, and limited cancer patterns in first- and second-degree relatives to determine eligibility for genetic testing. While both frameworks have been valuable in identifying LFS candidates, none of the probands in our study cohort met the Classic LFS criteria, highlighting potential limitations in real-world clinical settings. Consequently, our analysis focused exclusively on comparing LFSPRO performance against the Chompret criteria, which has become the more clinically relevant standard in contemporary practice.

**Table 1.**
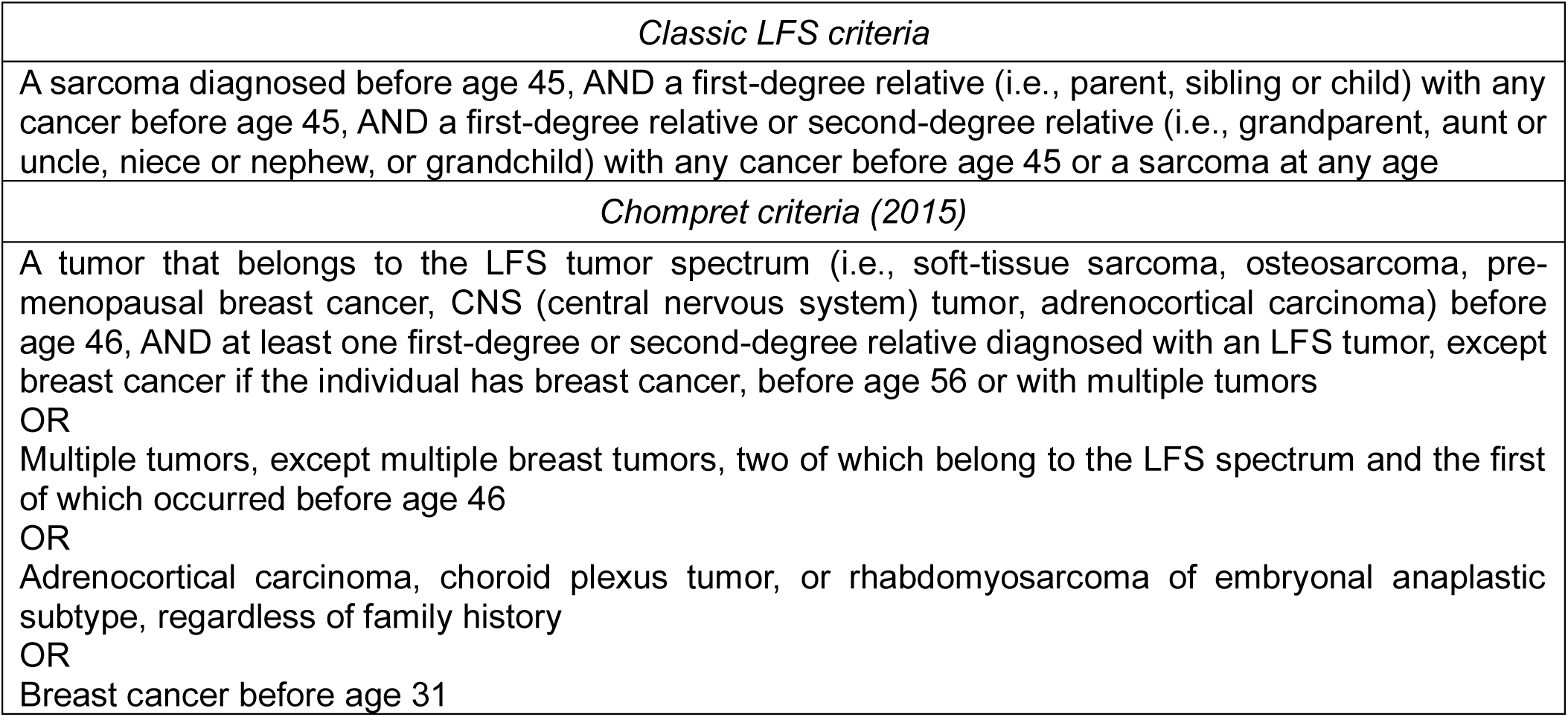
Description of Li-Fraumeni Syndrome Classic and Chompret Criteria.

### LFSPRO Statistical Framework

LFSPRO was developed as a Mendelian risk prediction model to estimate the probability that an individual carries a deleterious *TP53* mutation based on detailed family cancer history^15^. The model adopts a Bayesian formulation in which the carrier probability is obtained by updating the population prevalence with the likelihood of the observed pedigree and cancer diagnoses:

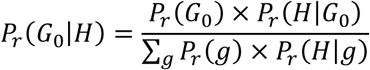

Here, *G*_0_ denotes the genotype of the counselee, *H* represents the family cancer history, and *g* ∈ {0,1} corresponds to the possible genotype states (wildtype or mutation carrier, including both heterozygous and homozygous mutation genotypes). The prior probability *P*_*r*_(*G*_0_) is set to the population prevalence of *TP53* mutations (0.0006)^15^, following the Hardy-Weinberg equilibrium for homozygotes. The likelihood term *P*_*r*_ (*H*|*G*_0_) represents the probability of observing the family cancer history given the counselee’s genotype, updated from integrating over all possible genotype configurations of relatives while incorporating Mendelian inheritance and age-specific cancer penetrance. Because the number of genotype configurations grows exponentially with pedigree size, LFSPRO employs the Elston-Stewart or peeling algorithm^20^, which recursively factors the pedigree into anterior (ancestral) and posterior (descendant) components conditional on a pivot individual, and marginalizes over unobserved genotypes as individuals are peeled off. This recursion enables efficient likelihood evaluation in extended pedigrees without enumerating all possible genotype states. Importantly, LFSPRO also models de novo *TP53* mutations, which arise spontaneously in the germline even when both parents are noncarriers. Such events account for an estimated 7-20%^21^ of LFS cases and are therefore clinically significant. In the model, they are represented by a transmission probability of 2*α*, where *α* is the per-allele mutation rate. By incorporating both inherited and de novo events, LFSPRO yields more accurate carrier probability estimates across diverse pedigrees.

Building on the original formulation, the model has been extended to support prospective cancer risk prediction. The Cancer-Specific (CS) model^22^ employs a competing risk approach with cause-specific hazard functions to estimate the probability of different cancer types as the first primary tumor, and the Multiple Primary Cancer (MPC) model^23^ uses recurrent event models based on non-homogeneous Poisson processes to capture the elevated risk of subsequent malignancies. By providing individualized, quantitative risk estimates rather than binary classifications, LFSPRO and its extensions enable more refined genetic counseling compared with traditional clinical criteria. Validation across multiple independent cohorts has demonstrated superior sensitivity and specificity for identifying *TP53* mutation carriers^16,17,24^, underscoring its clinical utility in LFS family risk assessment and management.

### Clinical Implementation

These statistical models are integrated into LFSPROShiny^18^, an interactive R/Shiny application that provides genetic counselors and healthcare providers with a user-friendly interface for inputting patient information and generating individualized risk assessments.

For this study, GCs utilized LFSPROShiny to facilitate clinical implementation. The interface allows GCs to efficiently input patient information from MDACC electronic health records to compute the carrier probabilities and generate individualized risk assessments. While GCs can select between the CS and MPC models for specific cases, the MPC model was set as the default based on its better performance compared to the CS model in prior validation studies^15,17^. Furthermore, we used a binary classification threshold of 0.2 for LFSPRO risk scores, as established in prior work^15^, where probands scoring ≥0.2 were classified as screening positive for potential *TP53* LP/P variant carrier status. This threshold achieved good trade-offs between sensitivity and specificity on multiple LFS datasets^15^.

### Survey Design and Administration

To evaluate GCs’ experience using LFSPRO in clinical practice, we developed a structured REDCap (Research Electronic Data Capture)^20,21^ survey instrument addressing four key domains: (1) concordance between LFSPRO predictions and standard clinical assessment, (2) additive value of LFSPRO beyond conventional genetic counseling, (3) comfort level with communicating LFSPRO results to patients, and (4) practical applications of LFSPRO-derived risk predictions in patient counseling. GCs taking these surveys could select to complete a survey for 1) a patient undergoing *TP53* germline testing, 2) a patient with a *TP53* germline variant that wanted to utilize LFSPRO’s cancer risk prediction, or 3) a combination of *TP53* germline testing, risk assessment, and cancer risk prediction. Each survey contained both structured quantitative measures and open-ended response fields for GCs to provide further feedback on their use of LFSPRO.

### Statistical Analysis

The performance of LFSPRO and Chompret criteria was evaluated using multiple complementary approaches. Diagnostic accuracy was assessed through sensitivity, specificity, positive predictive value (PPV), and negative predictive value (NPV) with 95% confidence intervals. Sensitivity measured the proportion of true *TP53* mutation carriers correctly identified by each model, while specificity measured the proportion of individuals without *TP53* mutations correctly identified as negative. PPV assessed how often a positive prediction was correct, and NPV assessed how often a negative prediction was correct, providing measures of clinical reliability for each prediction outcome. Discriminatory ability was quantified using receiver operating characteristic (ROC) curve analysis, with area under the curve (AUC) values and 95% confidence intervals calculated for both LFSPRO and Chompret criteria. An AUC of 0.5 represents no discriminatory ability (equivalent to random chance), while 1.0 represents perfect discrimination.

Statistical significance of associations was assessed using two-sides Fisher’s exact test, which was selected due to the limited sample size with cell counts below 5 in some contingency table cells. This non-parametric test was used to evaluate: (1) the association between model-based classifications (LFSPRO and Chompret criteria) and actual *TP53* mutation status, and (2) the association between prediction methods and GCs’ clinical judgment. The null hypothesis in each case was that no association exists between the compared variables, with a *p*-value < 0.05 considered statistically significant. Confusion matrices were visualized as heatmaps to illustrate the distribution of true positives, false positives, true negatives, and false negatives for each prediction method. For the GC survey data, descriptive statistics were calculated as percentages for categorical variables.

## Results

### Cohort Description

From December 2021 to March 2025, a total of 178 probands were included in this concordance study. The cohort comprised 115 (64.6%) assigned female at birth and 63 (35.4%) assigned male at birth, with a mean age of 30 years. Among the 148 probands who completed *TP53* genetic testing, 23 (15.5%) tested positive for a LP/P *TP53* germline variant, including one with a mosaic variant identified in two separate tissue types, and one with low variant allele frequency (VAF) suggestive of possible mosaicism. The majority (n=125, 84.5%) tested negative for germline *TP53* variants, though this group included one individual with a *TP53* variant at 13% VAF that could not be detected in a second tissue source, and another with a variant of uncertain significance in *TP53* that was likely mosaic. Four individuals had genetic testing but with results still pending, while 26 probands had no genetic testing results because they either declined testing or remained undecided. Since the Bayesian Mendelian model used in LFSPRO does not account for mosaicism, probands with possible mosaic *TP53* variants were excluded from all downstream analysis. See **Table 2** for a comprehensive summary of proband characteristics stratified by genetic testing results.

**Table 2.**
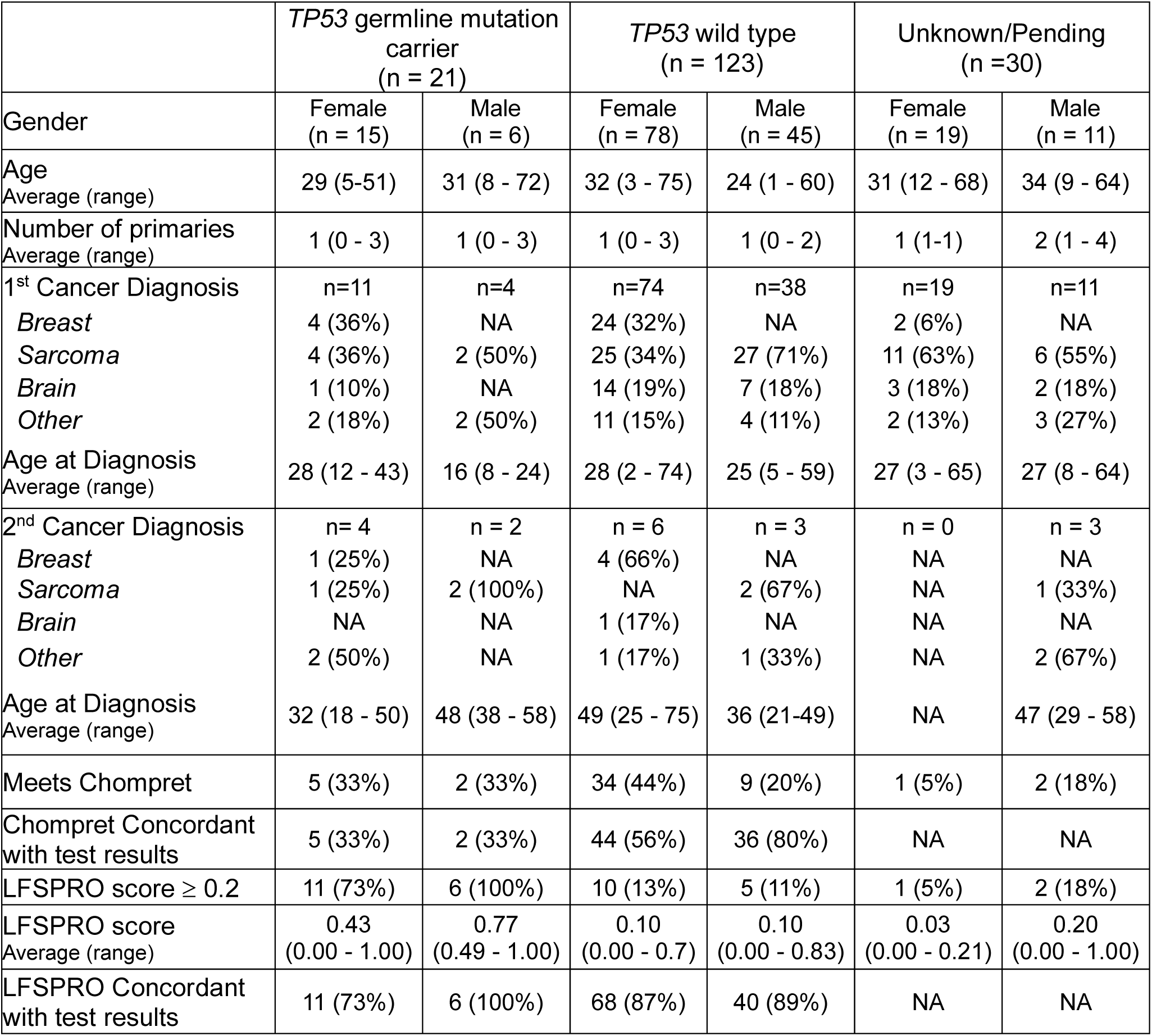
Summary statistics of prospectively collected probands concerning for LFS determined by GCs.

This prospectively collected clinical cohort represents a unique dataset of individuals flagged by GCs as concerning for LFS in real-world practice. The 15.5% mutation detection rate and inclusion of complex cases with mosaicism and uncertain variants provides an ideal testing set for evaluating LFSPRO’s clinical utility.

### LFSPRO Outperforms Existing Criteria

Among individuals with germline LP/P *TP53* variants (n=21), 15 (71.4%) had at least one primary cancer and 7 (28.6%) had at least two primary cancers. Of these individuals, only 7 (33%) met Chompret criteria for genetic testing, while 17 (81.0%) were identified by LFSPRO with a risk score (carrier probability) ≥ 0.2. In the mutation-negative group (n=123), 112 (91.1%) had at least one primary cancer and 9 (7.3%) had at least two primary cancers. Within this group, 43 (35.0%) met Chompret criteria while only 15 (12.2%) had an LFSPRO risk score ≥ 0.2. Alternatively, 80 (65.0%) individuals did not meet Chompret criteria, and 108 (87.8%) had LFSPRO scores below the 0.2 threshold. See **Fig. 2a** for detailed confusion matrices. Also, a two-sided Fisher’s exact test analysis confirmed LFSPRO’s statistically significant concordance with genetic testing results (*p*<0.001), whereas Chompret criteria failed to demonstrate significant association (*p*=1.00). A corresponding Sankey diagram (**Fig. 2b)** further illustrates the classification flow of probands by LFSPRO and Chompret criteria against genetic testing results.

**Figure 2.**
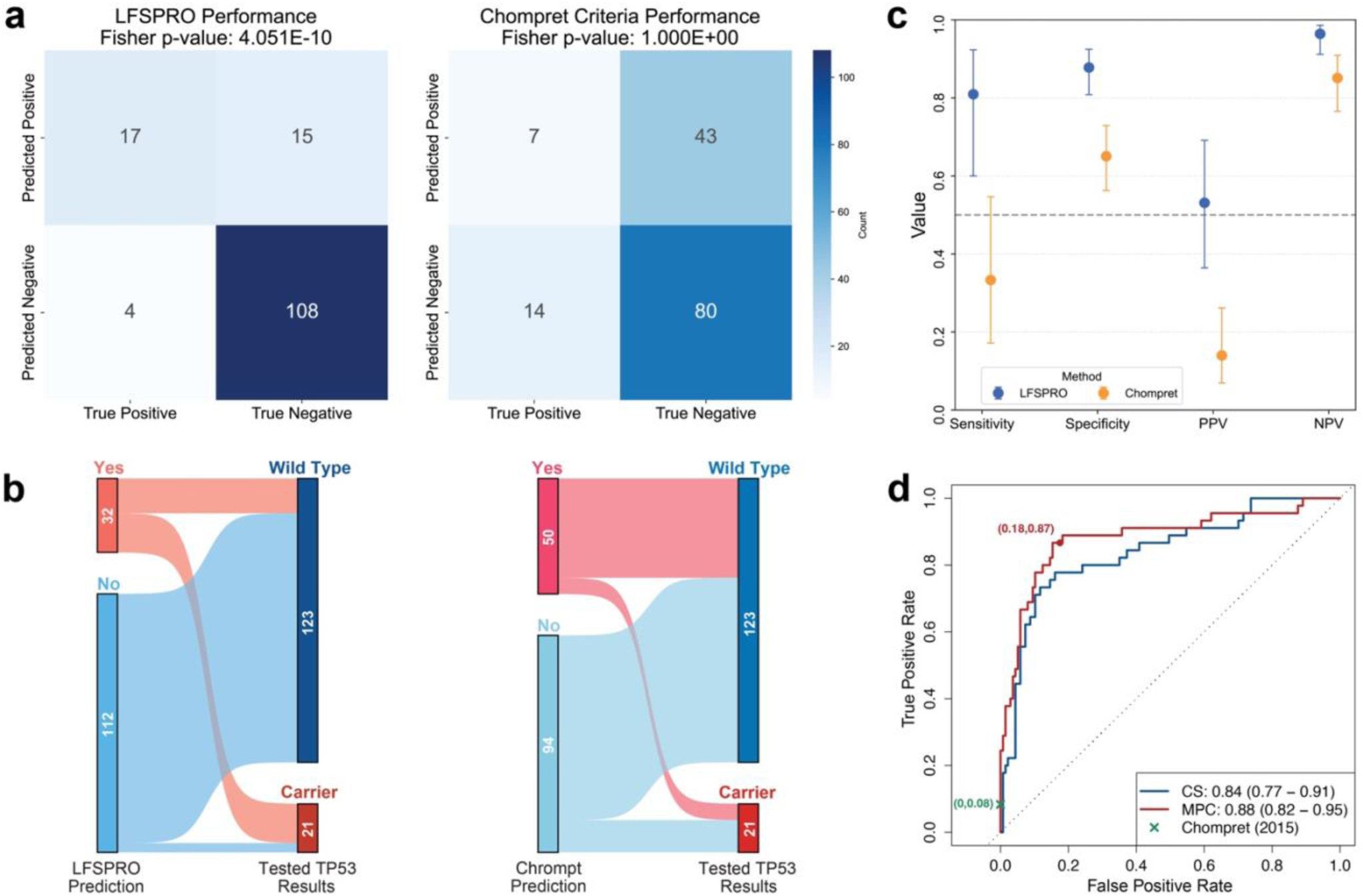
Comparative analysis of LFSPRO and Chompret criteria performance. **(a)** Confusion matrices for LFSPRO and Chompret criteria with associated Fisher’s exact test p-values. **(b)** Sankey diagrams showing the flow relationships between prediction methods and genetic testing results. **(c)** Performance metric comparison between LFSPRO and Chompret criteria across sensitivity, specificity, positive predictive value (PPV), and negative predictive value (NPV). **(d)** ROC curves comparing discriminatory performance of LFSPRO models and Chompret criteria. The red point indicates the MPC model’s performance at the 0.2 threshold.

LFSPRO demonstrated significantly better discriminatory performance over the conventional Chompret criteria for identifying *TP53* LP/P mutation carriers. LFSPRO achieved higher sensitivity of 0.810 (95% CI: 0.600-0.923) compared to Chompret’s 0.333 (95% CI: 0.172-0.546), and better specificity of 0.878 (95% CI: 0.809-0.925) versus 0.650 (95% CI: 0.563-0.730), as illustrated in **Fig. 2c**. Importantly, LFSPRO demonstrated substantially higher PPV at 0.531 (95% CI: 0.364-0.691) compared to Chompret’s 0.140 (95% CI: 0.070-0.262), indicating greater clinical utility for correctly identifying true mutation carriers. Similarly, LFSPRO’s NPV of 0.964 (95% CI: 0.912-0.986) outperformed Chompret’s 0.851 (95% CI: 0.765-0.909), providing greater confidence in ruling out *TP53* mutations when classified as negative. ROC curve as shown in **Fig. 2d** further demonstrated LFSPRO’s discriminatory ability with AUC scores of 0.88 (95% CI: 0.82-0.95) for MPC model, and 0.84 (95% CI: 0.77-0.91) for CS model. Calibration analyses using observed-to-expected (O/E) ratios further supported model validity. The MPC model, which was used as the default for carrier probability estimation based on prior validation studies, demonstrated good calibration (O/E 1.07, 95% CI: 0.80-1.34), with close agreement between predicted and observed carrier frequencies. The CS model showed modest underestimation of carrier probabilities (O/E 1.46, 95% CI: 1.09-1.84).

These findings demonstrate that LFSPRO outperforms Chompret criteria in identifying individuals with *TP53* LP/P variants, showing enhanced ability to predict true positives while reducing false positives, and strong alignment with genetic testing results. This suggests LFSPRO’s potential to assist in clinical decision-making and assist patients’ decisions on genetic testing.

### Genetic Counselors Report Positive Experience with LFSPRO

Beyond comparing LFSPRO with established clinical criteria, we evaluated its concordance with GC’s clinical judgment, the actual decision-making standard in practice. Four experienced GCs completed 64 surveys after counseling sessions. None of the probands in these sessions met Classic LFS criteria and only 20.3% met Chompret criteria. In practice, however, genetic counselors recommended germline testing for 77% of probands, underscoring that current criteria fail to capture many individuals whom clinicians consider appropriate for testing. Within these cases, LFSPRO provided additional decision support in 16%, highlighting its potential to complement clinical judgment when guideline frameworks alone are insufficient. Furthermore, GCs indicated they were comfortable sharing LFSPRO results with patients in 75% of cases, most often providing generalized summaries (39.1%) or specific risk estimates (35.9%), as shown in **Fig. 3a**.

**Figure 3.**
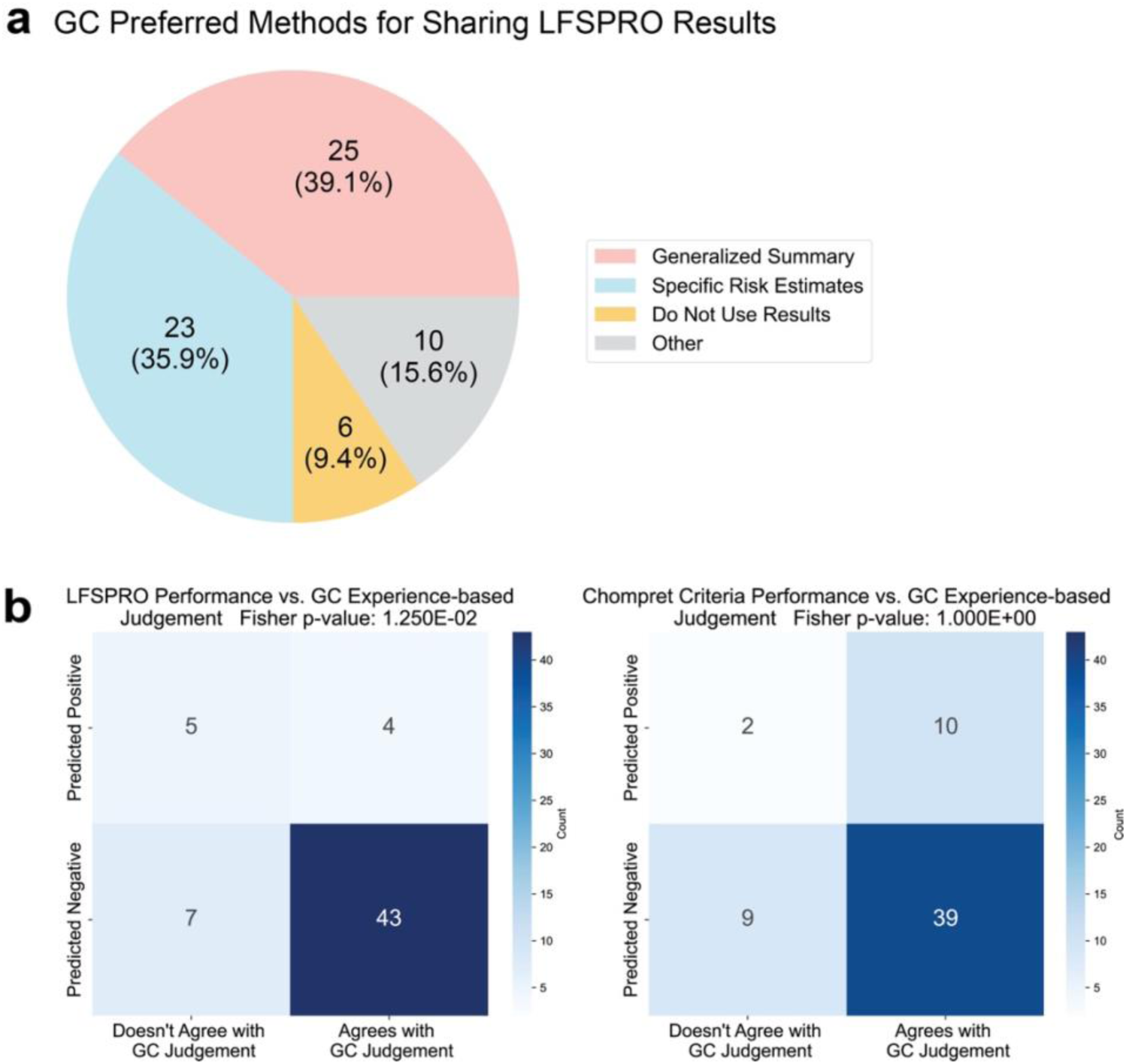
Statistical Analysis of GC Survey Results. **(a)** Genetic counselors’ preferred methods for communicating LFSPRO results to patients. **(b)** Contingency table analysis comparing between LFSPRO and Chompret Criteria with genetic counselor clinical judgment using two-sided Fisher’s exact test.

Fisher’s exact test demonstrated a significant association between LFSPRO predictions and GCs’ clinical judgment (p=0.01), with differential agreement rates when LFSPRO predicted above the cutoff (44.4%) versus below the cutoff (86.0%). In contrast, Chompret criteria showed no significant association with counselor intuition (p=1.0), yielding similar agreement rates regardless of whether criteria were met (**Fig. 3b**). Importantly, GC experience and clinical judgement were not incorporated as features in the LFSPRO model, making this concordance an independent validation of the model’s ability to capture clinically relevant risk patterns through objective variables alone. Together, these findings suggest that LFSPRO is usable, aligns more closely with clinical expertise than guideline criteria, and holds promise for integration into routine practice to support personalized risk assessment in LFS. Additional qualitative feedback from free-response comments further highlighted counselors’ perspectives on LFSPRO’s clinical implementation, with representative quotations presented in **Table 3**.

**Table 3.** Genetic counselor feedback on LFSPRO clinical utility by clinical scenario.

| Clinical Scenario | Genetic Counselors Quotes |
| --- | --- |
| Patient anxiety considerations | Patient was highly anxious about cancer history given her and her full siblings have all developed cancer. I think having a risk model would be helpful, but I worry that giving her exact numbers would have made her anxious and/or hyperfocus on those risk estimates. But if this was a less anxious patient, I would probably feel comfortable giving exact risk estimates. |
| Validating clinical judgement | Patient with uterine rhabdomyosarcoma and no other family history, I felt like the LFSPRO risk was accurate with my gut instinct. This family really wanted specific risk numbers (ex. they wanted specific numbers regarding how often RMS is genetic, they didn't feel like the general 5-10% of cancers are hereditary was specific enough for them). So I think this family would have really liked a personalized risk model where I could have provided the patient with risks tailored to her. |
|  | LFSPRO would have helped me better counsel this family- they came in thinking they likely had LFS, and I provided reassurance that they likely don't, but wasn't able to give them a percentage likelihood- 13% seems like what I would have quoted but LFSPRO's prediction would have validated my gut and made me more comfortable sharing a percentage with them |
| Supporting testing decisions | Family is undecided on testing- may decide to test in the coming weeks; I think this risk prediction would have been helpful for the family, as they had assumed the risk was MUCH higher |
|  | This family was wanting to know the risk to test positive to make a decision on the risks versus benefits of testing- I shared my prediction of <5%, this would have been helpful to quote more specifically. |

## Discussion

This prospective study provides important and timely clinical validation of LFSPRO, a personalized cancer risk prediction tool for Li-Fraumeni Syndrome. Compared to the widely used Chompret and Classic criteria, LFSPRO demonstrated strong predictive performance, with significantly higher sensitivity, specificity, PPV, and NPV. AUC analysis further confirmed LFSPRO’s strong discriminatory ability, and its predictions were significantly concordant with genetic testing results, in contrast to Chompret criteria which showed no significant concordance. These findings position LFSPRO as a robust clinical tool for identifying individuals at risk for carrying LP/P *TP53* variants.

Chompret and Classic criteria have long guided genetic testing for LFS and are endorsed in the National Comprehensive Cancer Network (NCCN) guidelines. However, these criteria rely on specific and limited clinical features and can fail to capture individuals with atypical or limited family histories. As a result, patients who would benefit from testing may be overlooked, particularly when insurance coverage is tied to satisfying formal guideline criteria. LFSPRO potentially addresses this gap by generating individualized, probabilistic estimates of carrier mutation status and specific cancer risk based on detailed family and personal history, offering a more flexible and precise approach that may facilitate access to testing and lifelong screening protocols.

Beyond its diagnostic accuracy, this study assessed LFSPRO’s impact on clinical decision-making through 64 genetic counseling surveys. In the majority of cases, counselors reported the patient (77%) did not meet standard testing criteria but were still recommending testing based on clinical judgment, with LFSPRO providing additional decision support in 16% of these cases. Importantly, LFSPRO predictions showed significant concordance with GC clinical judgment while current standard criteria did not. These findings underscore LFSPRO’s potential to complement clinical expertise and facilitate more personalized genetic counseling, particularly in cases that fall outside established testing frameworks.

LFSPRO also offers longitudinal cancer risk estimates at 5-, 10-, and 15-year intervals, enabling GCs and other providers to deliver more personalized risk counseling. These individualized projections can help patients better anticipate their likelihood of carrying a *TP53* mutation or developing cancer, and may assist in making decisions about genetic testing and surveillance. Personalized risk communication has been shown to improve patient understanding and promote informed decision-making^25^. In related clinical contexts, more accurate risk estimates have also been associated with reduced anxiety and cancer worry, particularly in patients with high baseline anxiety^26^. We expect that LFSPRO has similar potential to improve not only clinical accuracy but also the patient experience in LFS counseling.

### Study Limitations

Our study highlights the improved sensitivity and specificity of LFSPRO over existing clinical testing guidelines; however, several limitations remain. LFSPRO produced both false-positive and false-negative predictions. In particular, 3 of the 15 false positives involved unaffected family members being test for a known familial *TP53* variant. LFSPRO assigned a ∼50% risk based on Mendelian inheritance, though these individuals tested negative. These cases may inflate the false-positive rate and underestimate the model’s true performance in diagnostic testing contexts. Additionally, one of the false positives had a reported familial LP/P variant in other hereditary cancer genes (NF1), likely contributing to a strong family history misattributed to *TP53* by LFSPRO. Among the four false negatives, one individual carried the c.638G>A founder mutation, which is associated with reduced cancer risk. Finally, LFSPRO relies on patient-reported personal and family history collected in clinical settings. Its accuracy may be limited in cases with incomplete family histories or when a de novo *TP53* mutation occurs in an individual without prior cancer or sufficient family history. These limitations are shared by current clinical testing guidelines and highlight opportunities for further refinement of LFSPRO to improve its utility in diverse clinical scenarios.

### Future Directions

This study demonstrates that genetic counselors feel comfortable using LFSPRO in clinical practice and find it particularly helpful for guiding testing decisions in families not meeting standard criteria. To broaden its impact, future work should evaluate its usability among non-genetics providers, particularly in community or resource-limited settings where expertise in LFS may be scarce, and incorporate structured training to support implementation. The results presented in this work, including LFSPRO’s strong performance compared to existing criteria and alignment with clinical judgement, may also help encourage patients, their family members, and providers to participate in future clinical trials that incorporate LFSPRO.

In parallel, methodological advances continue to expand the underlying statistical framework. A new unified Bayesian semi-parametric model jointly accommodates recurrent and competing risks and extends applicability to broader cancer survivor populations^27^. Future efforts will focus on validating this model in large independent cohorts and integrating it into LFSPRO, thereby enhancing its clinical utility across diverse settings.

### Conclusions

In summary, this prospective study demonstrates LFSPRO’s real-world clinical accuracy in identifying *TP53* mutation carriers and strong alignment with GCs’ clinical judgment. The tool’s potential integration into current standard clinical practice can potentially facilitate GCs’ work by providing personalized risk estimates that enhance decision-making and support effective communication with patients and family, particularly in complex cases where traditional criteria provide limited guidance.

## Data Availability

All data produced in the present study are available upon reasonable request to the authors

## Data and code availability

The LFSPRO software is publicly available on GitHub at https://github.com/wwylab/LFSPRO. The LFSPROShiny interactive application used in this study is open-source and available at https://github.com/wwylab/LFSPRO-ShinyApp, with a live version hosted at https://namhnguyen.shinyapps.io/lfspro-shinyapp-master/.

De-identified aggregate data supporting the findings of this study are available from the corresponding authors upon reasonable request and subject to institutional review board approval. Genetic counselor survey data and analysis script are available from the corresponding authors upon reasonable request, with participant identifiers removed to protect confidentiality.

## Acknowledgements

This study was supported by grants from Cancer Prevention and Research Institute of Texas (RP200383) and the National Institutes of Health (R01CA239342, P30CA016672).

## Author contribution

Conceptualization, B.K.A and W.W.; methodology, software, N.H.N., P.G., and W.W.; data curation, investigation, J.L.C., J.C., and A.W.; resources, A.G., and B.K.A., formal analysis, J.L.C., R.L., and E.B.D.; writing-original draft, J.L.C., and E.B.D; writing-review & editing, J.L.C., R.L., B.K.A., and W.W., visualization, R.L., supervision, B.K.A., and W.W., funding acquisition, B.K.A., and W.W.

## Web resources

National Comprehensive Cancer Network (NCCN) Guidelines: https://www.nccn.org/guidelines/

