## Supplemental Material for "Performance of LFSPRO *TP53* germline carrier risk predictions compared to standard genetic counseling practice on prospectively collected probands"

**Supplemental Materials**

LFSPRO Concordance Survey

Thank you for your interest in participating in the survey to determine the concordance of LFSPRO results to standard genetic counseling of individuals at risk of Li-Fruameni syndrome.  This survey was developed by Wenyi Wang’s lab, in the Department of Bioinformatics and Computational Biology, to gauge a genetic counselor’s perception of the concordance of LFSPRO’s risk prediction results to standard information provided to individuals during a genetic counseling session that includes discussion of risk for Li-Fraumeni syndrome. As a genetic counselor, you are being asked to complete this survey to determine if the LFSPRO results from a particular counselee match with your professional judgement and if you would be comfortable in sharing such results with counselees in the future.   The data from this survey will be used to determine the baseline of concordance of LFSPRO to genetic counseling sessions and to potentially improve the functionality of LFSPRO. The survey will be closed after TBD surveys have been collected.   This survey should take less than 5 minutes to complete.

*Genetic Counseling Session Focus*

1. Please upload the LFSPRO results file. (100MB max file size)
2. What was the focus of your counseling session?

- TP53 genetic testing
- LFS cancer risk information
- Both TP53 genetic testing and cancer risk information

*TP53 germline testing*

1. Does the proband in this family satisfy the following TP53 germline testing criteria?

| **Criteria** | **Yes** | **No** |
| --- | --- | --- |
| Classic |  |  |
| Chompret |  |  |

2. On a scale of strongly disagree to strongly agree, please indicate your response to the following statement: As a genetic counselor, my intuition agrees with the current standard criteria for TP53 testing for this proband.

| **Strongly Disagree (1)** | **Disagree (2)** | **Undecided (3)** | **Agree**  **(4)** | **Strongly agree (5)** | **NA** |
| --- | --- | --- | --- | --- | --- |

3. What was your initial recommendation for testing?

- Single Gene TP53 testing
- Panel-testing including TP53 gene
- No testing

4. Did the proband elect for mutation testing to be performed?

- Yes
- No

5. Is the proband’s LFSPRO carrier risk probability above the 0.2 cutoff?

- Yes
- No

6. On a scale of strongly disagree to strongly agree, please indicate your response to each statement after reviewing the LFSPRO mutation carrier results:

| **Statement** | **Strongly disagree** | **Somewhat agree** | **Neither agree nor disagree** | **Somewhat agree** | **Strongly agree** |
| --- | --- | --- | --- | --- | --- |
| The mutation carrier prediction results match up to my gut expectation of this patient’s risk |  |  |  |  |  |
| I would have felt comfortable sharing the mutation carrier risk probability with the proband |  |  |  |  |  |
| After reviewing the LFSPRO results, I would have made a different recommendation regarding genetic testing |  |  |  |  |  |

7. If available, how would you have discussed the LFSPRO carrier risk results with this proband?

- “I would have provided the specific LFSPRO risk estimates.”
- "I would have provided a generalized summary of LFSPRO risk estimates."
- "I would have used the LFSPRO risk estimates to subtly guide or tailor the risk discussion without quoting or summarizing the proband’s LFSPRO risk estimates."
- "I would not have used the information provided by LFSPRO during this session."

8. Comments on TP53 germline testing prediction for this patient: ________
